# Neoadjuvant chemotherapy and adjuvant chemotherapy are similarly beneficial for five-year overall survival in locally advanced gastric cancer patients

**DOI:** 10.1101/2023.05.14.23289971

**Authors:** Sah Birendra Kumar, Yu Zhenjia, Lu Sheng, Zheng Yanan, Zhu Zhenglun, Li Jian, Li Chen, Yan Min, Zhu Zhenggang

**Author notes:** Birendra Kumar Sah, MD/Ph D Department of General Surgery, Gastrointestinal Surgery Unit Ruijin Hospital Shanghai Jiao Tong University School of Medicine Postal add: 197 Ruijin Er Road, Shanghai-200025, China Contact no. Yu Zhenjia MD/Ph D (Co-first author) Department of General Surgery, Gastrointestinal Surgery Unit Ruijin Hospital Shanghai Jiao Tong University School of Medicine Shanghai, China. Lu Sheng MD Department of General Surgery, Gastrointestinal Surgery Unit Ruijin Hospital Shanghai Jiao Tong University School of Medicine Shanghai, China. Zheng Yanan MD/Ph D Department of General Surgery, Gastrointestinal Surgery Unit Ruijin Hospital Shanghai Jiao Tong University School of Medicine Shanghai, China. Zhu Zhenglun MD/Ph D Department of General Surgery, Gastrointestinal Surgery Unit Ruijin Hospital Shanghai Jiao Tong University School of Medicine Shanghai, China. Li Jian MD/PhD Clinical Research Center Ruijin Hospital, Shanghai Jiao Tong University School of Medicine. Li Chen, MD/Ph D Department of General Surgery, Gastrointestinal Surgery Unit Ruijin Hospital Shanghai Jiao Tong University School of Medicine Shanghai, China. Yan Min, MD Department of General Surgery, Gastrointestinal Surgery Unit Ruijin Hospital Shanghai Jiao Tong University School of Medicine Shanghai, China. Prof. Zhu Zhenggang, MD/Ph D, FACS Department of General Surgery, Gastrointestinal Surgery Unit Ruijin Hospital Shanghai Jiao Tong University School of Medicine Shanghai Key Laboratory of Gastric Neoplasms Shanghai Institute of Digestive Surgery Shanghai, China. These authors contributed equally (Sah Birendra Kumar, Yu Zhenjia).

## Abstract

**Background:** Different types of neoadjuvant chemotherapy regimens have been compared for gastric cancer, mostly in terms of radiological downgrading or pathological tumor regression; however, no large-scale multicenter RCT study has conducted a head-to-head comparison of the overall survival rate between perioperative or neoadjuvant chemotherapy(NAC) and postoperative or adjuvant chemotherapy(AC). We explored whether the five-year overall survival rate was greater in patients who received perioperative chemotherapy plus surgery than in those who underwent surgery first and then had postoperative chemotherapy.

**Methods:** Altogether, 77 patients with a clinical diagnosis of cTNM stage III were included. Five-year overall survival rates (OS) were compared between patients who underwent neoadjuvant chemotherapy plus surgery (NAC) and patients who underwent surgery first plus adjuvant chemotherapy (AC). Propensity score matching was applied to adjust for the disparity between the two groups. A Kaplan‒Meier plot was created for survival analysis, and the log rank method was used to compare the difference in OS.

**Results:** A total of 34 patients were in the NAC group, and 43 patients were in the AC group. There was no significant difference in age (median 64 vs. 66 years), cTNM staging, or extent of gastrectomy between the two groups (p<0.05).The median follow-up time was 58 months (range of 53-65 months). The five-year overall survival (OS) rates for the patients in the NAC group and AC group were 61.8% and 73.5%, respectively. There was no significant difference between the two groups in the five-year overall survival rates (p>0.05). There was no significant difference in the severity grading of postoperative complications between the two groups (p>0.05).

**Conclusions:** There was no significant difference in the five-year overall survival rate between the patients who underwent perioperative chemotherapy plus surgery and those who underwent surgery first plus postoperative chemotherapy. A well-controlled prospective study is necessary to reconfirm whether perioperative chemotherapy is superior to postoperative chemotherapy for gastric cancer patients.

## Introduction

The treatment strategy for gastric cancer includes surgery and adjuvant therapy. In terms of survival benefits, the extent of surgery is no longer the focus of new research, and there are clear suggestions that chemotherapy is beneficial for locally advanced diseases. After the publication of the results from the MAGIC trial by David Cunningham in 2006, a new trend started for research on neoadjuvant chemotherapy (1). However, it should be noted that the MAGIC trial was the comparison between a group of patients who underwent perioperative chemotherapy (preoperative and postoperative chemotherapy) plus surgery with a group of patients who had surgery only, i.e., a group of patients who did not receive chemotherapy at all (1). In 2014, Sung Hoon Noh published the results of another famous trial, the CLASSIC trial, in which they compared a group of patients who had surgery only with a group of patients who had surgery and received postoperative chemotherapy (2). The 5-year overall survival rate of the patients who received perioperative chemotherapy plus surgery was significantly higher than that of the patients who underwent surgery only in the MAGIC trial. The five-year OS was significantly higher in the patients who underwent surgery and postoperative chemotherapy than in those who underwent surgery only in the CLASSIC trial. Despite the demographic differences between the two trials, these results at the very least suggest that chemotherapy was beneficial for gastric cancer patients because the 5-year OS was increased in the patients who received either perioperative chemotherapy or postoperative chemotherapy (1, 2). There was no large-scale multicenter RCT study, in which a head-to-head comparison has been conducted between perioperative chemotherapy and postoperative chemotherapy. Only recently, a multicenter RCT from China, the RESOLVE trial, published their results in the Lancet and compared perioperative chemotherapy with postoperative chemotherapy, but the patients in the two groups received different chemotherapy regimens (3). This study demonstrated that in patients who underwent surgical treatment, the 3-year DFS was significantly better in patients who received perioperative chemotherapy with the SOX regimen than in those who received postoperative chemotherapy with the CAPOX regimen (59.4% versus 51.1%).The 3-year DFS for the patients who received postoperative chemotherapy with the SOX regimen was 56.5% (3). However, it is still not clear whether perioperative SOX is also superior to postoperative SOX.

In addition, there are still blunt disagreements between Eastern and Western researchers over the indication of neoadjuvant chemotherapy (4-7). One reasonable argument is that preoperative evaluation of the tumor (cT and cN) is not consistent with pathological pT and pN. More than 10% of stage I patients were misdiagnosed with late-stage disease in a large-scale Japanese study (8). We explored whether the five-year overall survival rate was greater in patients who received perioperative chemotherapy plus surgery than in those who underwent surgery first and then postoperative chemotherapy.

## Methods

Only pathologically confirmed gastric cancer patients who were preoperatively diagnosed with cTNM stage III (cT3 or T4 and N1 or N2 or N3 and M0) and underwent radical surgery (total or partial gastrectomy with D2 lymphadenectomy) were included in this study. All patients were treated in 2018 at Ruijin Hospital, Shanghai Jiaotong University School of Medicine, which is a referral center for gastric cancer patients in China. Patients were divided into two groups, perioperative chemotherapy plus surgery group (NAC) and surgery first plus chemotherapy group (AC). Because of the retrospective nature of the study, we empirically excluded 403 patients for different reasons that we considered may adversely affect the analysis (Supplementary Table 1). Our previous study for elderly patients suggested that there was a difference in overall survival between the patients who had total gastrectomy and the patients who had partial gastrectomy; therefore, we applied the propensity score matching method to eliminate the discrepancy between the two groups. Altogether, 172 cases were included in the propensity score matching for cT staging, cN staging, and types of gastrectomy. Among them, 98 cases were matched and approached for follow-up. Finally, only 77 patients were included in the final analysis, and 21 patients were excluded due to insufficient or unconfirmed data for survival time. Postoperative morbidity and mortality were recorded according to Clavien‒Dindo grading (9).The overall survival (OS) time in this study is the time from the date of surgery to death from any cause. The data for relapse-free survival (RFS) were not accurately available and thus were not analyzed in this study.

## Statistical analysis

The statistical analysis was performed with Statistical Package for Social Science (SPSS) version 22.0 for Windows (SPSS, Inc., Chicago, Illinois). Nonparametric methods were used to analyze data with an abnormal distribution. The continuous data are expressed as the median and range, and the Mann‒Whitney U test was used for continuous data. The chi-square test and Fisher’s exact test were used to compare the differences between the two groups as appropriate. Survival data were presented as the length of overall survival (OS) in months. A Kaplan‒Meier plot was created for survival analysis, and the log rank method was used to compare the OS rates. Cox regression analysis was used to identify the risk factors for OS. A p value of less than 0.05 was considered statistically significant.

## Results

Altogether, 77 patients were included in this study:34 patients in the NAC group and 43 patients in the AC group. There was no significant difference in demographic variables between the patients of the two groups (p>0.05), including the preoperative clinical staging of the gastric cancer (cT stage and cN stage) and the extent of gastrectomy (Table 1). Patients received the established chemotherapy regimens; among 34 patients in the NAC group, 13 patients received SOX, 9 patients received EOX, 7 patients received FLOT, and 5 patients received SOX-A. Patients in both groups received postoperative chemotherapy for six months, and there was no significant difference in the completion of postoperative adjuvant chemotherapy between the two groups (p>0.05). Details on the adverse effects of chemotherapy were not described due to the lack of accurate data due to the retrospective nature of the current study. The median follow-up time was 58 months (range of 53-65 months). The estimated five-year overall survival (OS) rates for the patients in the NAC group and AC group were 61.8% and 73.5%, respectively. There was no significant difference between the two groups in the five-year overall survival rates (p>0.05). There was no significant difference in the severity grading of postoperative complications (Table 2) or length of postoperative stay between the two groups (p>0.05).One patient in the NAC group achieved pCR, and there were 9 patients in the AC group with pTNM stage I (Table 3).

**Table 1.** Demographic Data.

| Parameter |  | NAC | DS | P Value |
| --- | --- | --- | --- | --- |
| Sex | Male | 26(76.5) | 26(60.5) | 0.106 |
|  | Female | 8(23.5) | 17(39.5) |  |
| Age (years) | Median(Range) | 64(38-77) | 66(44-86) | 0.531 |
| Body mass index | Median | 22.42 | 22.18 | 0.573 |
| Site of tumor | Proximal | 8(23.5) | 9(20.9) | 0.227* |
|  | Body | 11(32.4) | 7(16.3) |  |
|  | Distal | 15(44.1) | 25(58.1) |  |
|  | Total | 0 | 2(4.7) |  |
| cT stage | T3 | 3(8.8) | 4(9.3) | 1.000 |
|  | T4 | 30(88.2) | 37(86.2) |  |
|  | T4b | 1(2.9) | 2(4.7) |  |
| cN stage | N1 | 2(5.9) | 1(2.3) | 0.709 |
|  | N2 | 22(64.7) | 27(62.8) |  |
|  | N3 | 10(29.4) | 15(34.9) |  |
| Type of Gastrectomy | Distal | 17(50.0) | 23(53.5) | 0.821 |
|  | Total | 17(50.0) | 20(46.5) |  |
| Postop Chemo | Yes | 26(76.5) | 31(72.1) | 0.307 |
|  | No | 5(14.7) | 11(25.6) |  |
|  | Unknown | 3(8.8) | 1(2.3) |  |

\*Fisher's Exact Test

**Table 2.** Postoperative Complications.

| Complication | NAC plus Surgery | Direct Surgery |
| --- | --- | --- |
| Grade 0- I | 27(79.4) | 21(72.4) |
| Grade II | 6(17.6) | 7(24.1) |
| Grade IIIa | 1(2.9) | 0 |
| Grade IVa | 0 | 1(3.4) |
| Grade V | 0 | 0 |

**Table 3.** Postoperative Pathology.

| Parameter | Stage | NAC plus Surgery | Direct Surgery |
| --- | --- | --- | --- |
| ypTNM | pCR | 1(2.9) |  |
|  | I | 3(8.8) |  |
|  | II | 11(32.4) |  |
|  | III | 19(55.9) |  |
| pTNM | I |  | 9(20.9) |
|  | II |  | 5(11.6) |
|  | III |  | 29(67.4) |

## Discussion

For the last three decades, it has been reported that more than 2300 articles have been published regarding neoadjuvant chemotherapy for gastric cancer (10); however, very few of them were widely accepted, and if there is only one article that should be remembered as a pioneering concept for neoadjuvant chemotherapy, it is the MAGIC trial (1), despite not being the first one to report. Despite exponential growth in terms of publications on neoadjuvant chemotherapy, most of these studies are mainly conducted in a few countries. China, the USA, and Japan scored as one of the top three countries for these studies (10). There is still no joint agreement or a strict guideline to recommend that neoadjuvant chemotherapy is mandatory for gastric cancer. This may be due to conflicting results from different centers (6,7,11-13) or perhaps somewhat deficient study designs in the past, and none of them answered the fundamental question that perioperative or neoadjuvant chemotherapy plus surgery is superior to surgery first plus adjuvant chemotherapy(1,2,3). Most of the research on neoadjuvant chemotherapy is focused on comparing the efficacy between different types of chemotherapy regimens, mainly focused on radiological or pathological responses to therapy (15, 16). Al-Batran SE. published the initial results of the FLOT 4 trial and demonstrated that the FLOT regimen was better than ECF or ECX in terms of pathological regression and tolerance (15). Sah BK published the initial reports of comparative results of pathological efficacy between the triplet chemotherapy FLOT and the doublet chemotherapy SOX. There was no significant difference between neoadjuvant FLOT and SOX in terms of tumor regression grading; however, the five-year overall survival rate has not yet been published (16). In addition, we did not find any study that claimed the superiority of NAC over AC in terms of overall survival. Even in the final results of the FLOT 4 trial in 2019, the five-year overall survival rate was much lower than that reported in the CLASSIC trial (2, 17). The main concern for neoadjuvant chemotherapy in Japan is the accuracy of clinical TNM staging (8), and it should be noted that 20.9% of patients in the surgery-first group were pTNM stage I in this study. Because it would have impacted the comparison of OS, we further compared the OS between the two groups excluding pathological stages I and II, although it was not required because a similar probability of misdiagnosis was also anticipated in the NAC group. Nonetheless, there was no difference in OS between the two groups even after excluding these early-stage tumors (Fig.2).

**Fig.1.**
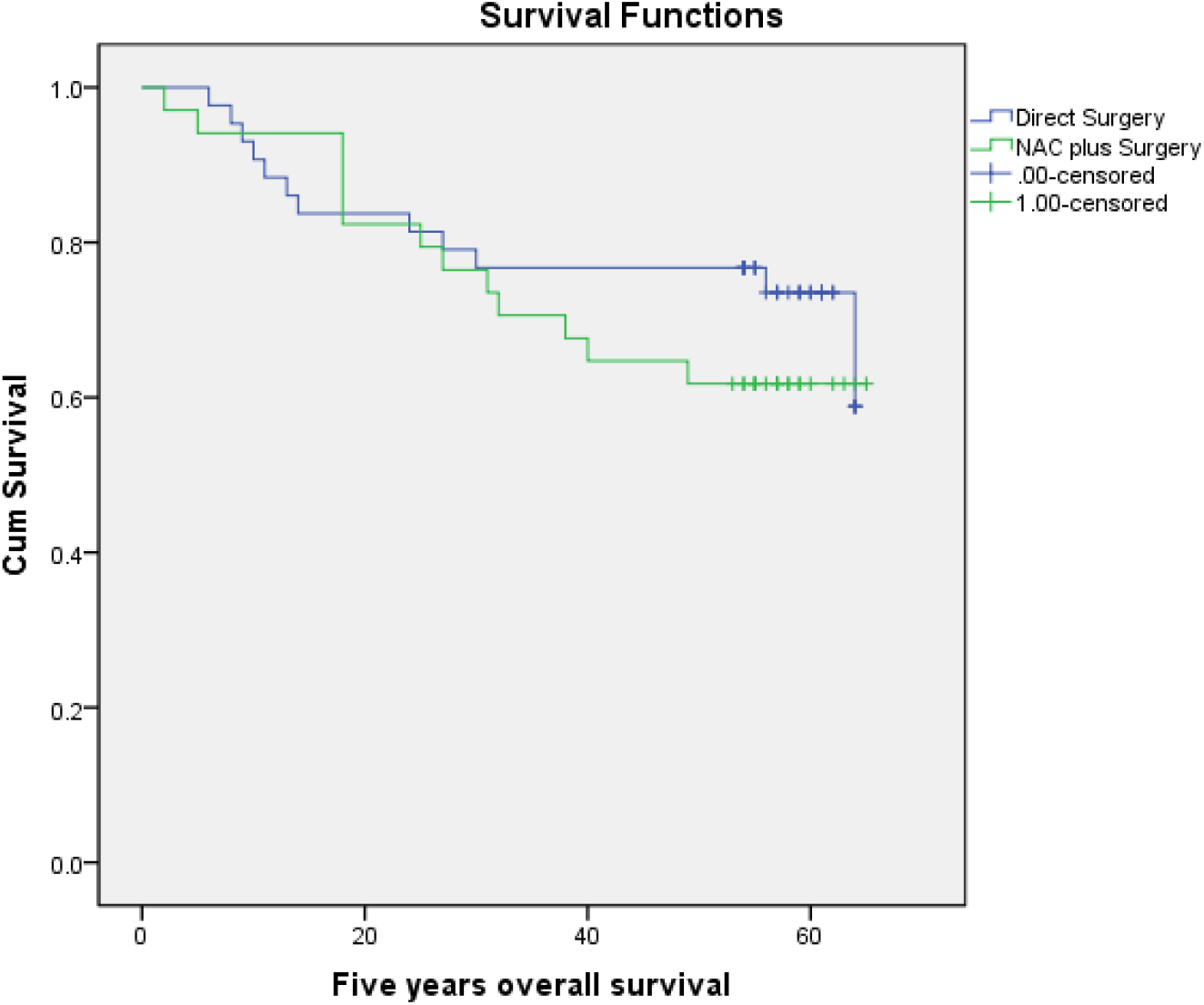
Kaplan-Meier (K-M) Plot for OS.

**Fig.2.**
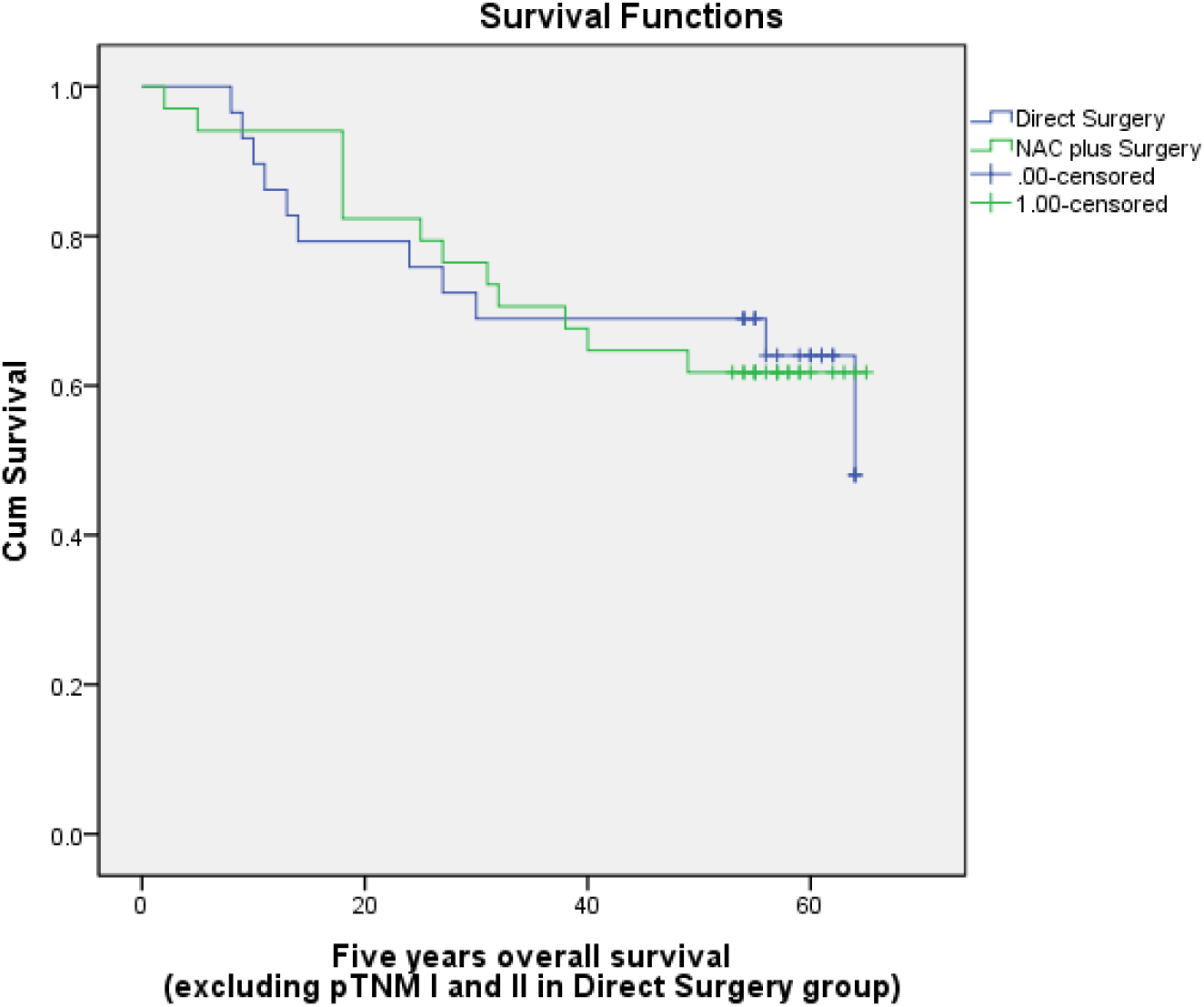
Kaplan-Meier (K-M) Plot for OS (only pTNM III in Direct Surgery group)

There are several shortcomings in this study, mainly due to retrospective analysis; the number of patients finally included in this study is too small to achieve any convincing conclusion. We excluded hundreds of patients for valid reasons (Supplementary Table 1) to eliminate the disparity between the two groups. There were no data on the adverse effects of chemotherapy; we did not analyze it because the focus of this study was to compare the overall survival rate, and many adverse effects were not recorded in standard forms. In addition, there were several types of chemotherapy regimens, although the types of chemotherapy might have somehow influenced the result, but previous studies have shown that the CAPOX and SOX regimens were similarly effective for gastric cancer (3). Even SOX was similarly effective as the FLOT regimen in terms of pathological regression (16). Nevertheless, despite being a small-scale retrospective study, the result of this study is quite interesting, and it raises the fundamental question of whether neoadjuvant chemotherapy is beneficial because the CLASSIC trial has shown that the 5-year overall survival rate is 78% for patients who underwent surgery first and then received postoperative or adjuvant chemotherapy (AC). Therefore, any rationale for alteration to this treatment should only be considered for two basic reasons, either to increase the percentage of overall survival rate or decrease the toxic side effects of chemotherapy. By the analysis of these data, we neither deny the use of neoadjuvant chemotherapy nor question its efficacy in terms of overall survival. However, at the very least, this study concludes that chemotherapy either perioperatively or postoperatively is similarly effective. Whether the addition of preoperative chemotherapy is beneficial in terms of OS is still questionable. There is still a place for a better-designed study to be conducted. Perhaps the next study should focus on the noninferiority test and not on the superiority test between the two modes. If NAC is similarly effective to AC, then it would be better to conduct a large-scale study that compares total preoperative chemotherapy plus surgery (no postoperative chemotherapy) with surgery first plus postoperative chemotherapy.

Despite the lack of concrete data to support this, we can easily assume that preoperative chemotherapy is better in terms of completion of required cycles compared to postoperative chemotherapy after major surgery. Due to postoperative complications, a certain number of patients cannot start adjuvant chemotherapy on time or are simply unable to receive chemotherapy.

The result of this study by no means negates the need for further study on neoadjuvant chemotherapy. However, attention is necessary for designing future studies by understanding the limitations of conventional chemotherapy. Whether the addition of immune therapy or targeted therapy is beneficial for long-term survival is still unknown.

## Conclusion

There was no significant difference in the five-year overall survival rate between the patients who underwent perioperative chemotherapy plus surgery and those who underwent surgery first plus postoperative chemotherapy. A well-controlled prospective study is necessary to reconfirm whether perioperative chemotherapy is superior to postoperative chemotherapy for gastric cancer patients.

## Data Availability

The datasets used and/or analyzed during the current study are available from the corresponding author upon reasonable request.

## Declaration

### Ethics approval and consent to participate

The Ethics Committee of Ruijin Hospital approved this study and waived the individual informed consent due to the retrospective analysis. The study was carried out in conformity with the Declaration of Helsinki (as revised in 2013)

### Consent for publication

The manuscript does not contain any individual data which identifies the patient included in this study.

### Patients and the public

Patients and the public were not involved in the design of this study due to the retrospective nature of this research.

### Author contributions

BKS designed the study, collected the patient data, and drafted the manuscript.ZJY followed up with patients for overall survival and revised the manuscript. SL assisted in data collection from the central database of the unit.YNZ and ZLZ revised the drafts of the manuscript.CL and ZGZ participated in the design of the study and critically revised the drafts of the manuscript. All authors meet the criteria for publication; all authors have read and approved the final manuscript.

## Acknowledgments

The authors thank all the clinicians in the gastrointestinal department for their support to conduct this study. And especially thanks to Mrs. Qin Yu for recording the data in the central database.

---

**Supplementary Table 1.**

| Reason for exclusion | Number |
| --- | --- |
| No standard data for preoperative CT | 22 |
| Preoperative diagnosis of T1, T2, N0 | 192 |
| Clinical suspicion of M1 or MX | 48 |
| Combined multiorgan resection | 41 |
| Robotic surgery | 8 |
| Thoraco-abdominal approach for the surgery | 3 |
| Surgery for remnant gastric cancer | 2 |
| Preoperative chemotherapy intended for conversion therapy | 2 |
| Unconventional chemotherapies | 12 |
| Neuroendocrine carcinoma | 6 |
| Squamous cell carcinoma | 2 |
| No R0 surgery | 10 |
| Surgery performed by inexperienced surgeons(<30surgeries) | 55 |

